# Patterns and Determinants of Outcomes in Cervical Spine Injury Patients: A Retrospective Study at AaBET Hospital, Addis Ababa, Ethiopia

**DOI:** 10.1101/2024.08.29.24312801

**Authors:** Alemayehu Beharu Tekle, Nikodimos Eshetu Dabe, Molla Asnake Kebede, Ayalew Zewde Tadesse, Bisrat Solomon Zewge, Melaku Tsediew Berhanu

## Abstract

A cervical spinal cord injury is a tragic occurrence for the sufferer and their loved ones. Because so many resources are needed to manage the patient during both the acute and rehabilitative stages, it has a significant impact on society and it is mainly related to cervical spine fractures and the most frequent kind of spinal fractures are those to the cervical spine. Automobile accidents, followed by diving into shallow water, firearm injuries, and sports activities are common causes of cervical spine injury. In developing countries like Ethiopia, little is known about the prevalence of cervical spinal injuries. An understanding of the prevalence of spinal injury is fundamental to developing possible preventive strategies and improving our primary trauma care. Assessing the pattern, outcome, and associated factors of patients with cervical spine injury who have visited AaBET Hospital from January 1, 2018 to November 30, 2023 is the primary objective of this study. Institution-based cross-sectional study was conducted at AaBET Hospital, Addis Ababa, Ethiopia. The study included patients who presented to the emergency department with a diagnosis of cervical spine injury from January 1, 2018, to November 30, 2023. Descriptive analysis was used for statistical analysis of baseline data, and regression analysis was used to determine associations between dependent and independent variables. A p-value <0.05 was considered statistically significant. A total data of 149 patients were analyzed, with an average age of 36.3 ± 14.9 years (ranges 9-85 years) and the male-to-female ratio was 2.9:1. Road traffic accident occurs in 49.7% of patients as a mechanism of injury followed by falling down accident (39.6%) of patients. Seventy-eight (52.3%) patients sustained with a total of 114 associated injuries (ASOI). Head injury was the commonly associated injury followed by chest and extremity injury. The most frequently injured cervical vertebra was C7 followed by C6 and T1. 68.5% of the patients have multilevel injuries. 33.6% of patients have neurological impairment ASIA class A followed by ASIA class E (29.5%). The overall hospital mortality is 7.4%. The level of cervical spine injury, the ASIA class of the patient, and the presence of associated injury were strongly associated with mortality. The mean ± SD length of hospital stay was 13.6 ± 16.4 days. And 30.2% of patients have prolonged lengths of hospital stay (PLOS). Neurosurgical intervention and the presence of associated injury have been significantly associated with PLOS. This study showed the common mechanism was RTA and C7 was the common injury level. C3 injury level, ASIA A neurologic deficit, and having associated injury were associated with mortality. Undergoing neurosurgical intervention and the presence of associated injury were associated with prolonged length of hospital stay (PLOS).

## Introduction

Severe neurological abnormalities following spinal cord injury (SCI) frequently result in long-term disability. Because the diaphragm and intercostal muscles’ innervation is disrupted in cervical SCI, respiratory function is worse, necessitating long-term mechanical ventilation. Given that using mechanical ventilation raises the risk of respiratory issues (1–3).

SCIs (spinal cord injuries) are a serious public health risk. A cervical spinal cord injury is a tragic occurrence for the sufferer and their loved ones. Because so many resources are needed to manage the patient during both the acute and rehabilitative stages, it has a significant impact on society and it is mainly related to cervical spine fracture (4,5). The most frequent kinds of spinal fractures are those to the cervical spine, which are frequently linked to SCI (6). According to a 1996 study, the incidence rate of spine fractures at all sites was 64 per 100,000, while the rate of cervical fractures was estimated to be 12 per 100,000 based on a population survey conducted in Canada (7).

Even though most cervical spine injuries are very modest, severe trauma can cause physical impairment and reliance that lasts a lifetime. Depending on local differences, auto accidents, and sports-related incidents are the leading causes of cervical spine injuries. Factors such as age, gender, and the mechanism of damage can affect both the severity and the result of the injury (8–10).

The most frequently overlooked serious injuries are those to the cervical spine (C-spine), which can have disastrous effects on the patient and have significant medico-legal ramifications for the surgeon performing the surgery or the emergency room (11). Due to research on cervical spine injuries being limited in Ethiopia, this study intends to describe the patterns and outcomes of patients with cervical spine injuries who have visited AaBET Hospital, affiliated with St. Paul’s Hospital Millennium Medical College Emergency Department, in Addis Ababa, Ethiopia, from January 1, 2018 to November 30, 2023. As a result, the primary goal of this study is to evaluate the trends, results, and contributing variables of patients with cervical spine injuries who visited AaBET Hospital in Addis Ababa, Ethiopia, between January 1, 2018 and November 30, 2023.

## Methods and Materials

### Study Area/Setting and Period

An institution-based cross-sectional study was conducted from April 1, 2023, to November 30, 2023, at AaBET Hospital in Arada Sub-city, Addis Ababa, Ethiopia. AaBET Hospital was inaugurated as a public hospital under St. Paul’s Hospital Millennium Medical College (SPHMMC) in 2015 and is one of the first health facilities with a dedicated trauma and burn unit. The hospital includes departments of Emergency Medicine and Critical Care (EMCC), Neurosurgery, Orthopedics and Traumatology, and an academic program in Plastic and Reconstructive Surgery. It has a well-structured emergency and ICU setup with more than 50 beds in the Emergency Department (ED), organized into red, orange, yellow, and green zones, and includes 11 ICU beds and 3 semi-ICU beds. The hospital provides 24/7 services with comprehensive laboratory and radiography units, offering ultrasound, X-ray, and CT scan services.

### Study Population

The source population has included the records of all patients with spine injuries and who have visited AaBET Hospital from January 1, 2018 to November 30, 2023. The study population comprised all records of patients with cervical spine (C-spine) injuries and those who have visited AaBET Hospital from January 1, 2018 to November 30, 2023. The data of these clients were accessed from this hospital record and documentation room from April 1, 2023 to November 30, 2023.

### Eligibility

#### Inclusion Criteria and Exclusion Criteria

All patients with C-spine injuries who have visited AaBET Hospital from January 1, 2018 to November 30, 2023 were included. Patients with incomplete documentation, unclear information, or lost records were excluded from the study.

### Sample Size Determination and Sampling Technique

#### Sample Size Determination

The sample size was determined based on similar studies conducted at Tikur Anbessa Specialized Teaching Hospital (TASTH), which showed that 33% of patients with spinal cord injury had cervical spine injuries (12). The sample size was calculated using the single population proportion formula:

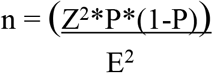

Where:

n = minimum sample size

P = estimated prevalence (33%)

α = level of significance (5%)

E = desired precision (5%)

Z = Z-score corresponding to 95% confidence interval (1.96)

Based on these parameters, the minimum sample size was calculated to be 339.7 and approximately 340. Given that the total number of patients with spinal cord injuries (the source population) visiting AaBET Hospital ED during the study period was limited to 410, the finite population correction formula was applied, resulting in a corrected minimum sample size of 185 patients. Since the total estimated number of patients with cervical spine injuries was close to this minimum, all eligible patients were included in the study.

Sample size correction formula, n=ni/ 1+ ((ni-1)/N)

n= corrected sample size, ni= Initial sample size, N=source population

n=340/1+(340-1)/420 = 185, but only 149 participants are included in this study as to the sampling techniques below.

#### Sampling Technique

A total of 410 patients with spine injuries were recorded in the registration book. Out of these, 30 charts were lost, and 9 charts were incomplete, leaving 371 complete charts. All patients with cervical spine injuries were included in these 371 charts (**Fig 1**).

**Fig 1.**
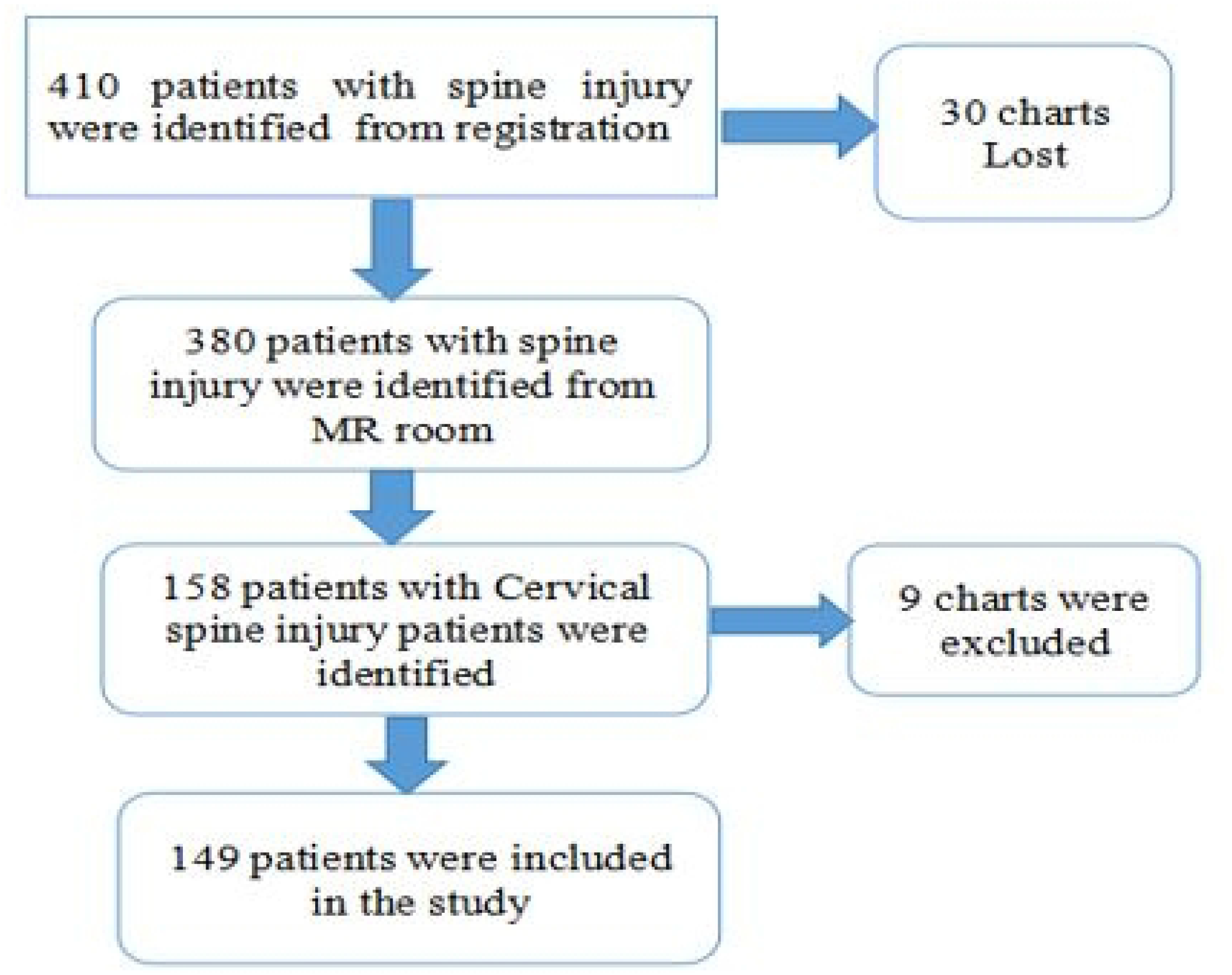
Schematic presentation of a sampling of study participants in AaBET Hospital, Addis Ababa, Ethiopia, 2023.

## Data Collection Method, Tools, and Procedures

Data were collected from the charts of all eligible cervical spine injury patients using a structured data extraction format. Nurses and interns from other hospital were recruited and trained as data collectors with principal investigators supervising the process. Data were manually cleaned and incomplete checklists were excluded from entry. Data were then entered into SPSS version 25 for analysis. The data collectors and supervisors didn’t have communication with the clients. The authors also didn’t have access to information that could identify individual participants during and after data collection.

### Study Variables

#### Independent Variables

Age, Sex, ED arrival time, Mechanism of trauma, Level of cervical spine injury, Type of cervical spine fracture, ASIA classification, Neurogenic shock, Associated injuries, Neurosurgical intervention

#### Dependent Variables

➢ Outcome at discharge (dead or alive)
➢ Hospital length of stay

### Operational Definitions

➢ **Pattern**: Distribution of C-spine injuries by sex, age, mechanism of injury, and residence among study participants (1).
➢ **Primary Outcome**: Condition of patients with C-spine injuries at discharge (dead or alive) (13).
➢ **Secondary Outcome:** Hospital length of stay(14).
➢ **Neurologic Improvement:** Improvement in ASIA classification (e.g., from ASIA A to ASIA B) (15).
➢ **Prolonged Hospital Stay (PLOS):** Hospital length of stay exceeding the 75^th^ percentile (14).

### Data Quality Control

The initial step in developing the data collection tool involved reviewing various English-language Publications and utilizing readily available resources(4,16–20). Prior to data collection, supervisors and data collectors were trained. The lead investigator checked the data extraction tool every day while data was being collected to ensure it was accurate, consistent, and comprehensive. The pre-test is typically administered to 5% of the sampled population, which is comparable to the study population at Menilik Comprehensive Specialty Hospital, two weeks before the actual data collection period. The instrument was reworded, made clearer, and organized differently in response to the pre-test results. After the pretest, a review was done to determine the questionnaires’ internal consistency and content validity. An appropriate amount of internal consistency was determined by looking at Cronbach’s alpha (> 0.7). To guarantee that the surveys were comprehensive during the actual data collection process, daily data entry and supervision were conducted.

### Data Processing and Analysis

A checklist was used to identify, gather, and verify the correctness of all patient charts pertaining to C-spine injuries who were seen at AaBET Hospital between January 1, 2018, and November 30, 2023. Epi Data statistical software version 4.6 was used to code, enter, clean, and verify the data. After that, SPSS version 27 was used to carry out the analysis. To look for missing values and variables, frequency and cross-tabulation were employed. The mean, percentage, frequency, and standard deviation of descriptive statistics were utilized to calculate the patients’ clinical and demographic data. A 95% confidence interval was utilized to determine the relationship between the dependent and independent variables using binary logistic regression. To take potential confounders into consideration, variables with p-values less than 0.25 were added to the multivariable model. To examine the degree of correlation between all independent and dependent variables, multivariate logistic regression was used; a p-value of less than 0.05 was deemed statistically significant.

### Ethical Considerations

This study used secondary data from patient charts, which were handled with strict confidentiality. Ethical clearance and approval to conduct the research were obtained from St. Paul Hospital Millennium Medical College and the Department of Emergency Medicine and Critical Care, Addis Ababa. The study commenced after obtaining ethical clearance and permission from the hospital management to review records. Due to the retrospective design, the requirement for informed consent was waived. The data collectors and supervisors were recruited from other government hospital and they didn’t have communication with the clients. The authors also didn’t have access to information that could identify individual participants during and after data collection.

## RESULTS

### The socio-demography Characteristics

The study group comprised 149 patients, including 111 males (74.5%) and 38 females (25.5%), resulting in an overall male-to-female ratio of 2.9:1. The mean age of the study population was 36.3 ± 14.9 years, with ages ranging from 9 to 85 years. The peak incidence occurred in the 21-30 age group. Most male patients sustained cervical spine injuries between 21 and 30 years of age, while female patients primarily exhibited injuries between 31 and 40 years (**Table 1**).

**Table 1:** Socio-demographic characteristics of the study participants in AaBET Hospital, Addis Ababa, Ethiopia, 2023 (n = 149).

| Variables | Category | Frequency | Percent |
| --- | --- | --- | --- |
| Age | ≤20 | 17 | 11.4 |
|  | 21-40 | 47 | 31.5 |
|  | 41-60 | 39 | 26.2 |
|  | >60 | 46 | 30.9 |
| Sex | Male | 111 | 74.5 |
|  | Female | 38 | 25.5 |
| Address of clients | Addis Ababa | 31 | 20.8 |
|  | Afar | 3 | 2.0 |
|  | Amhara | 23 | 15.4 |
|  | Gambella | 2 | 1.3 |
|  | Oromia | 73 | 49.0 |
|  | SNNPR | 15 | 10.1 |
|  | Somali | 1 | .7 |
|  | Tigray | 1 | .7 |
|  | Total | 149 | 100.0 |

### Emergency Department assessment

Among the 149 patients included in the study who were seen at the emergency department, 122 (81.9%) were referred cases from different regions of the country, while 27 (18.1%) were non-referral cases. The median time to arrive at the emergency department was 24 hours (IQR 44), with a range from 1 hour to 360 hours (15 days). Of the patients with cervical spine injuries, 107 (71.8%) were triaged to the yellow-green area, 18 (12.1%) to the red area, and 24 (16.1%) to the orange area. At presentation, 124 patients (83.2%) had neck pain or tenderness, 15 (10.1%) had no neck pain, and 10 (6.7%) were difficult to assess. Ninety-three patients (62.4%) arrived at the emergency department without a cervical collar, while 56 (37.6%) had a cervical collar in place. Among the 122 referred patients, only 56 (45.9%) arrived with a cervical collar, whereas 66 (54.1%) did not. Eleven patients (7.4%) presented with neurogenic shock, and eight patients (5.4%) with cervical spine injuries were intubated at the emergency department. Seven of them were intubated for airway protection due to severe traumatic brain injury and one for type 2 respiratory failure.

### Mechanism of trauma

Among the studied population, the most common cause of injury was road traffic accidents followed by falling accidents. Road traffic accidents were most common in the 21-40 age group, while falls were the predominant cause of injury in those over 60. Among the 59 patients who sustained injuries from falls, 47 (79.6%) fell from a height, 10 (17%) fell from a standing height, and 2 (3.4%) fell into a ditch (**Table 2**).

**Table 2:** Clinical pattern and outcome of patients with C-spine injury in AaBET Hospital, Addis Ababa, Ethiopia, 2023 (n = 149).

| Variables | Category | Frequency | Percent |
| --- | --- | --- | --- |
| Mechanism of trauma | RTA | 74 | 49.7 |
|  | FDA | 59 | 39.6 |
|  | Assault | 10 | 6.7 |
|  | A sharp object | 1 | .7 |
|  | Gunshot | 3 | 2.0 |
|  | Hit by a falling object | 1 | .7 |
|  | Other | 1 | .7 |
| ED disposition | Admitted to ward | 27 | 18.1 |
|  | Discharged from ED with a neurosurgery appointment. | 61 | 40.9 |
|  | Referred to other facility | 33 | 22.1 |
|  | Left against medical advice | 9 | 6 |
|  | Admitted to ICU | 12 | 8.1 |
|  | Died | 7 | 4.7 |
| ED length of stay in days | ≤1 | 3 | 2.0 |
|  | 1-7 | 100 | 67.1 |
|  | >7 | 46 | 30.9 |
| Total Length of hospital stay | <7 | 86 | 57.7 |
|  | 7-14 | 21 | 14.1 |
|  | 15-28 | 24 | 16.1 |
|  | 28-56 | 15 | 10.1 |
|  | >56 | 3 | 2.0 |
| Outcome at discharge | Dead | 11 | 7.4 |
|  | Alive | 138 | 92.6 |
| Hospital length of stay | N-LOS | 104 | 69.8 |
|  | E-LOS(PLOS) | 45 | 30.2 |

### Associated injury

Seventy-eight (52.3%) patients sustained a total of 114 associated injuries (ASOI). Among these, seventy patients (61.4%) sustained head injuries, twenty-five (21.9%) sustained chest injuries, fourteen (12.3%) sustained extremity injuries, four (3.5%) sustained pelvic injuries, and one (0.9%) sustained an abdominal injury. Of those who sustained head injuries, 51 (72.9%) had mild head injuries, 12 (17.1%) had moderate head injuries, and 7 (10%) had severe head injuries. Most of these associated injuries resulted from road traffic accidents and falls

### Injury characterization

#### Fracture localization

A total of 288 cervical spine fractures occurred in the 149 patients studied. The most frequently injured vertebra was cervical spine 7 (C7), accounting for 73 fractures (26%), followed by cervical spine 6 (C6) with 72 fractures (25.6%), and thoracic vertebra 1 (T1) with 53 fractures (18.9%).

#### Types of fracture

Most of the patients sustained burst fractures, accounting for 66 (28%) cases, followed by transverse process fractures in 64 (27.1%) patients, and spinous process fractures in 52 (22.0%) patients. Additionally, eight (3.4%) patients had hyper-extension teardrop fractures, four (1.7%) had unilateral facet dislocations, and three (1.3%) had cervical spinal cord injuries without associated cervical spine fractures. Approximately 18 (12.1%) patients with cervical spine injuries also had concurrent spine injuries at other sites. Specifically, fifteen (10.1%) patients had associated thoracic vertebrae fractures, and three (2%) patients had associated lumbar vertebrae fractures.

#### Neurological deficit

Among patients who sustained cervical spine injuries, 50 (33.6%) had neurological impairment classified as ASIA class A, while 44 (29.5%) were classified as ASIA class E. Additionally, 24 (16.1%) patients were classified as ASIA class C, 20 (13.4%) as ASIA class B, and 11 (7.4%) as ASIA class D (**Figure 2**).

**Figure 2:**
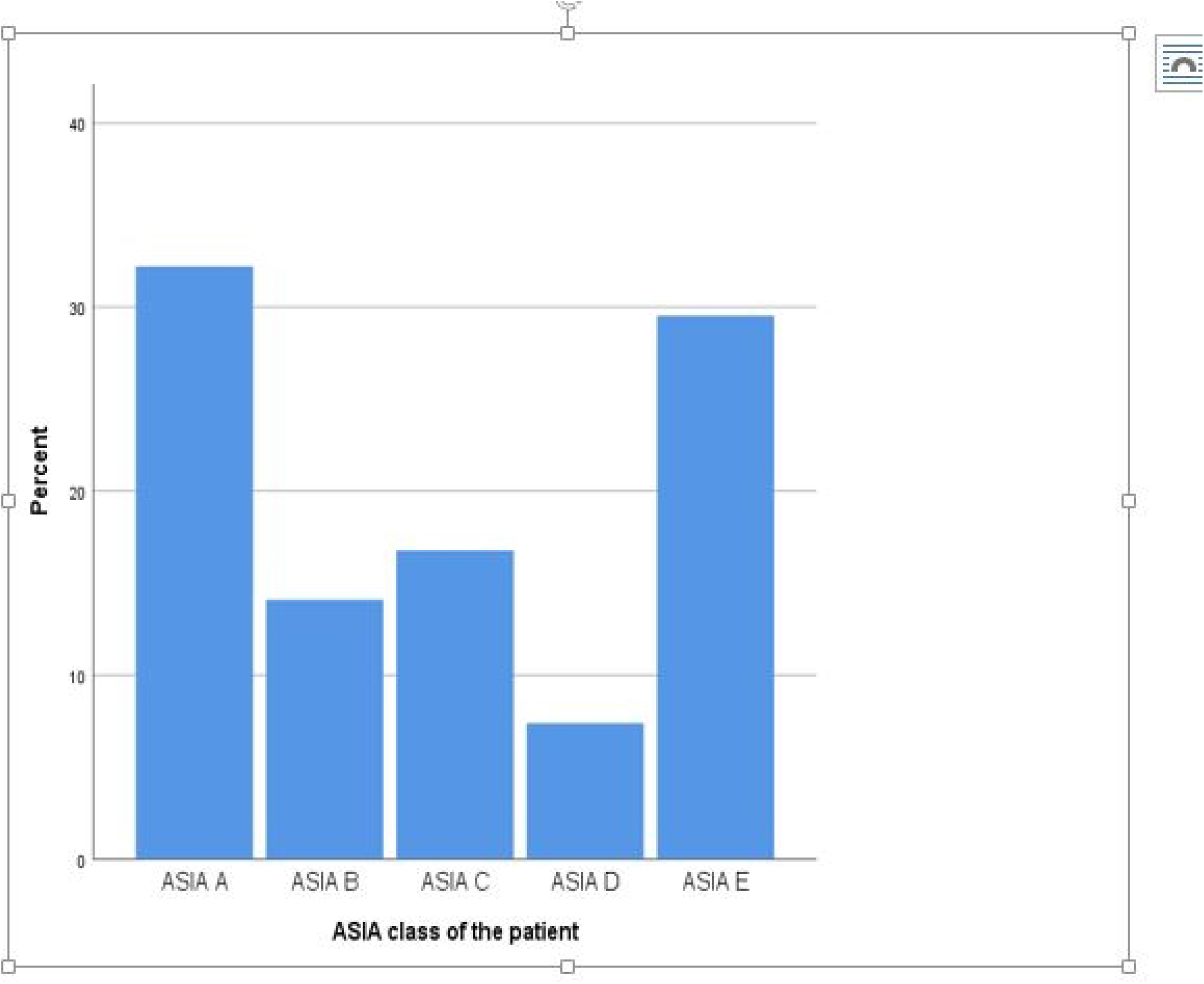
Neurologic deficit of the patients with c-spine injury in AaBET Hospital, Addis Ababa, Ethiopia, 2023 (n = 149).

#### Emergency Department (ED) disposition

Among the 149 patients seen at the ED, 27 (18.1%) were admitted to the ward, 61 (40.9%) were discharged from the ED with a neurosurgical appointment, 33 (22.1%) were referred to another facility, 9 (6%) left against medical advice (LAMA), 12 (8.1%) were admitted to the ICU, and 7 (4.7%) died at the ED. Among the 27 patients admitted to the ward, 23 underwent cervical spine surgery, while 4 patients underwent cervical spine surgery among those admitted to the ICU (**Table 2**).

Type 2 respiratory failure was identified as the cause of death in all patients who died at the ED. The median length of stay in the ED was 4 days (IQR 5), ranging from 1 to 31 days. Only 3 patients had stays of less than 24 hours, while the majority were discharged within one week; approximately 46 patients stayed in the ED for more than one week. The mean ± SD length of total hospital stay for the study population was 13.6 ± 16.4 days, with a minimum of one day and a maximum of 91 days. The 75^th^ percentile for hospital stay length was 15 days, indicating that 45 (30.2%) patients stayed longer than this percentile. Among the twelve patients admitted to the ICU, four died while the remainder were discharged alive, resulting in an overall hospital mortality rate of 7.4% (11/149), including the seven deaths at the ED.

#### Factors associated with mortality

In this study, all independent variables potentially influencing mortality were included in the regression model. To identify factors associated with mortality while adjusting for potential confounders, independent variables with a p-value ≤ 0.25 in binary logistic regression were included in a multiple regression model to calculate adjusted odds ratios, quantifying their association with the dependent variable. Variables such as age category, patient sex, level of cervical spine injury, ASIA class of the patient, presence of neurogenic shock, and presence of associated injuries were found to be associated with death in bivariate regression analysis. Following multivariable logistic analysis, only the level of cervical spine injury, ASIA class of the patient, and presence of associated injuries showed statistically significant associations with mortality (**Table 3**).

**Table 3:**
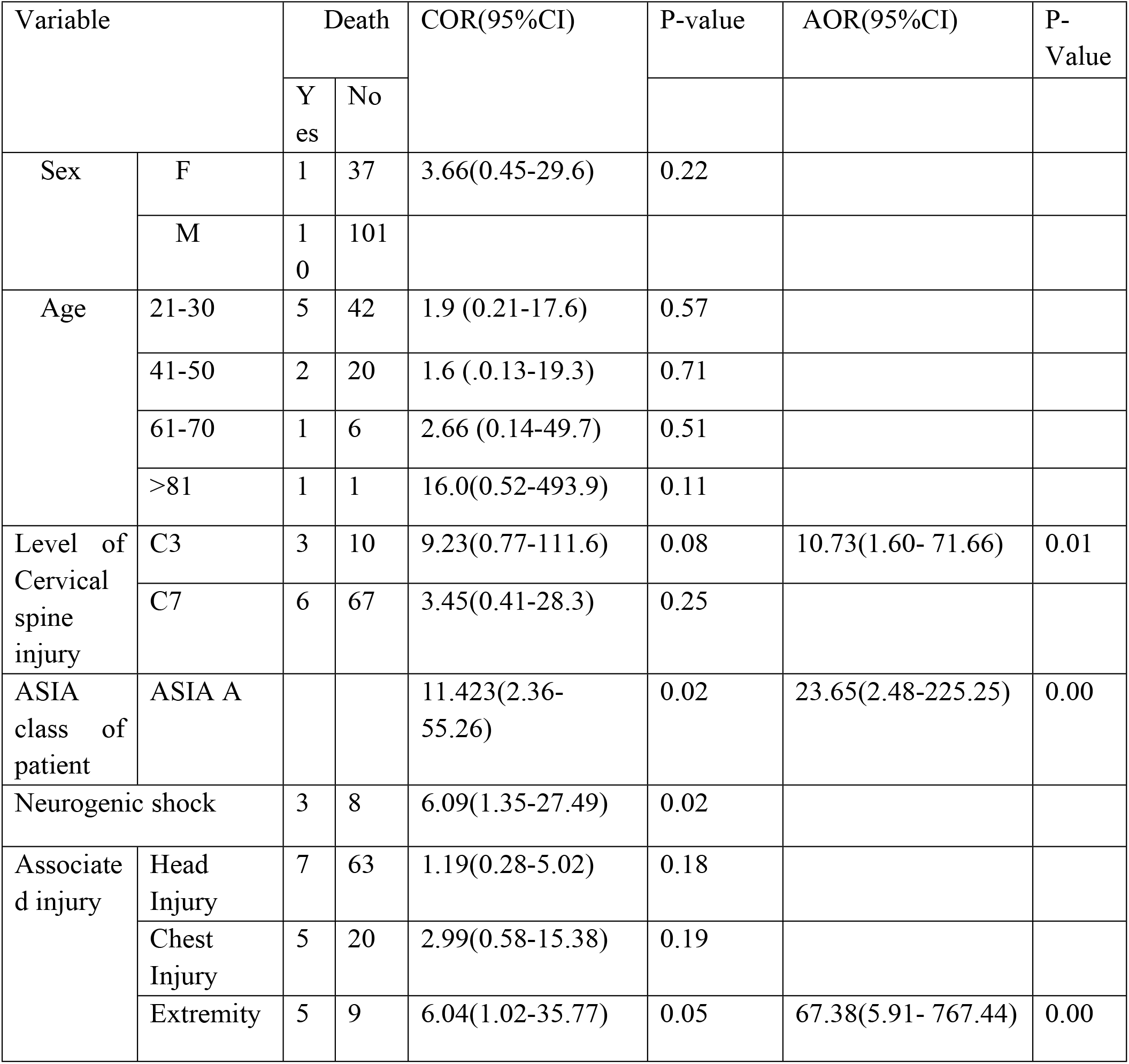
Bivariate and multivariate Analysis to identify factors associated with mortality in patients with cervical spine injury in AaBET Hospital, Addis Ababa, Ethiopia, 2023 (n = 149).

#### Factors associated with Long hospital stay

In this study, all independent variables potentially influencing prolonged hospital length of stay (PLOS) were included in the regression model. To identify factors associated with PLOS while controlling for possible confounders, independent variables with a p-value ≤ 0.25 in binary logistic regression were selected for inclusion in a multiple regression model. Adjusted odds ratios were then computed to quantify the strength of association with the dependent variable. Variables such as ASIA class of the patient, neurosurgical intervention, presence of associated injuries, and hospital-acquired infection were found to be associated with PLOS in bivariate regression analysis. Following multivariable logistic analysis, the variables that showed statistically significant associations with PLOS were neurosurgical intervention and the presence of associated injuries, particularly chest injuries, which were strongly associated with prolonged hospital stay (PLOS) (**Table 4**).

**Table 4:**
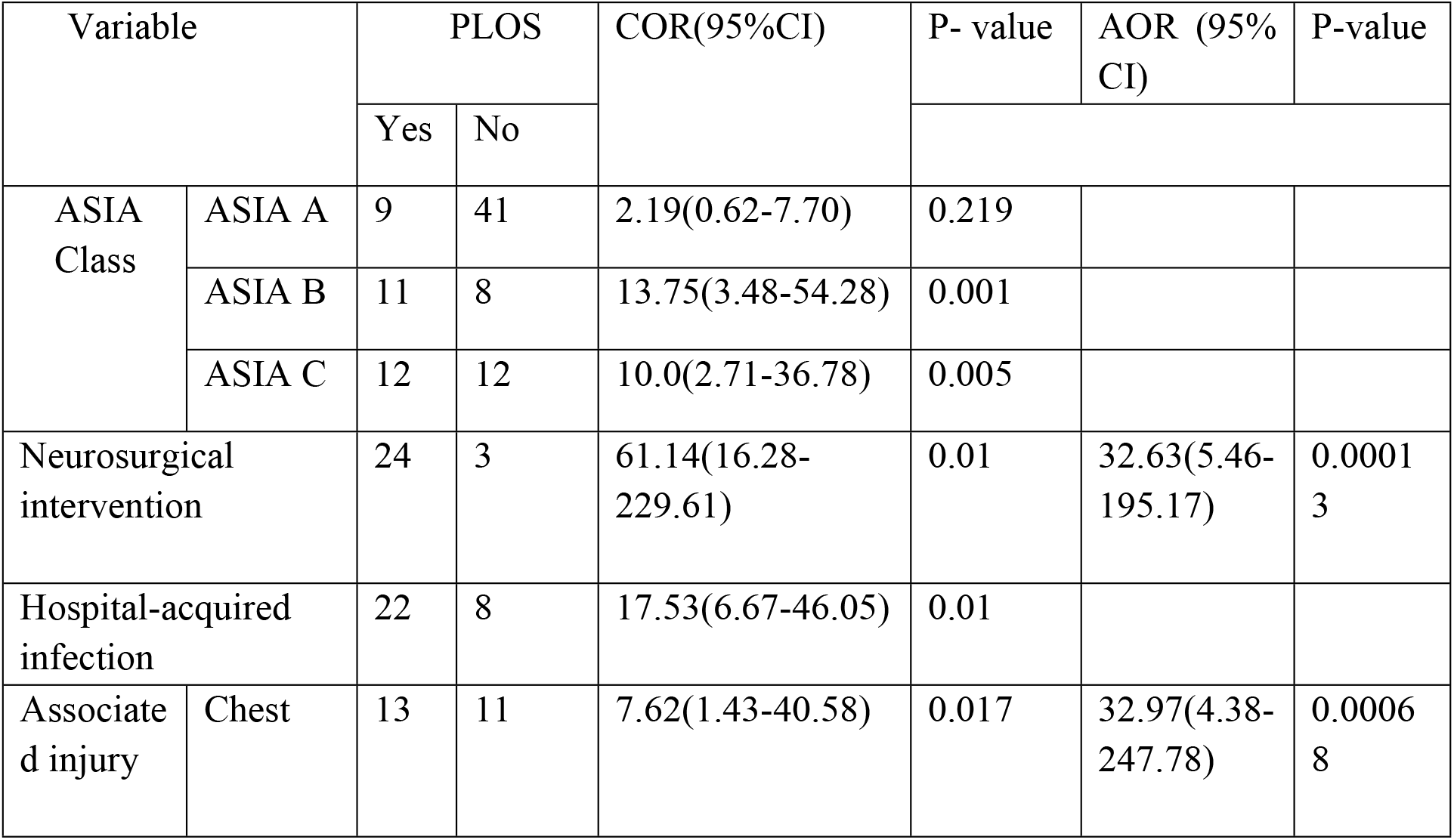
Bivariate Analysis to identify factors associated with Hospital PLOS in patients with cervical spine injury in AaBET Hospital, Addis Ababa, Ethiopia, 2023 (n = 149).

## Discussion

It is critical to comprehend the pattern of cervical spinal injuries and the variables linked to poor outcomes (death or extended hospital stays) in order to enhance primary trauma care and potentially create preventive measures.

Our study revealed that men are more likely than women to suffer from cervical spine injuries, which is consistent with earlier research (13,15,21). This is mainly due to men being more likely exposed to the major risk factors of spine injury (MVA) and While men in similar situations mostly experienced substantial injury to their cervical spines, women involved in auto accidents displayed statistically more serious lesions to their lumbar spines (22).

The mean age at the time of injury ranges from 32.8 years in Addis Ababa Black Lion Hospital to 34 years in Iran and 56 years in Norway (12,23,24). Our findings support the majority of literature, showing that the research population’s mean age was 36.3 years, with a peak age between 21 and 30 years, which is associated with greater active motor and vital activity.

Approximately 50% of all injuries worldwide are SCIs, and the majority of these occur in traffic incidents involving automobiles, bicycles, or pedestrians. According to a study done in Iran, falls are the second most common trauma that results in cervical spine injuries after motor vehicle crashes (23,25). Road traffic accidents were the most frequent cause of accidents among the population under study. This mechanism resulted in injuries for 74 patients (49.7%), with falls accounting for 59 (39.6%) of these cases. A small number of patients had gunshot and assault-related cervical spine injuries.

One common finding in this study sample is associated injuries. 114 associated injuries (ASOI) were sustained by 78 patients (52.3%). The most frequent connected injury is a head injury, which is followed by an extremity injury and a chest injury. This result is consistent with the findings by Prasad V. et al., which noted the existence of related injuries such as severe facial, head, and neck trauma (11). Additionally, according to Philipp Leucht et al., and other investigations, head, thoracic, and extremity injuries were the most often reported related injuries (26–29).

Cervical spine 7 (C 7) with 73 (26%), cervical spine 6 (C 6) with 72 (25.6%), and thoracic vertebra 1 (T1) with 53 (18.9%) fractures were the most commonly injured cervical vertebras. 102 patients, or 68.5%, have multiple injuries. According to a study by Higashi T and his colleagues, the majority of injuries happened at the levels of C4 (30%) and C5 (33%). The study also revealed that the injury level was dispersed from C3 to T1 (20).

It is believed that more than half of spinal cord injuries are incomplete (20,24), Similarly, according to our research, approximately 66.4% of the patients (99/149) had either no neurological damage at all or an incomplete injury. Merely 33.6% of patients suffering from cervical spinal cord injuries possessed an ASIA A scale.

Numerous studies have documented that patients with cervical spine cord injuries die at a rate ranging from 19% in a population research conducted in Norway to 63.9% in a retrospective multi-center study carried out in Germany involving patients with severe cervical spine injuries (30–33). Compared to the previous research, our study’s death rate is 7.4%. The observed variations in the clinico-demographic makeup of the population under study may account for a portion of the disparity. For example, the German study included patients with an abbreviated injury score (AIS) of 6 for severe cervical spine injuries, whereas the Norwegian study excluded patients with multiple site injuries and associated severe head injuries, which may increase the risk of death in patients with cervical spine injuries. The study population’s mean age was 80 years (range 65-101 years), whereas the current study’s mean age is 36.3 ± 14.9 years (range 9-85 years). Therefore, among other things, a younger age may be linked to a reduced death rate, in addition to that the majority of research indicated a strong correlation between aging and the chance of dying from traumatic SCI (31,33). The deceased group in this study was 50 years of age or younger as a result, aging is highly related to death.

Due to a neurological deficiency, cervical spinal cord injuries frequently result in severe, lifelong impairment. Numerous investigations have demonstrated a link between mortality and severe paralysis (19,20,31). Similarly, this study demonstrates that a significant risk factor for mortality in severe cervical SCI was an ASIA impairment scale of A. One review also found that paralysis of the muscles in the chest wall and abdomen can cause respiratory compromise, necessitating long-term mechanical ventilation. Complete paralysis can affect both muscle groups and result in severe breathing issues, especially in cases of lower cervical SCI (34).

There is debate on the relationship between mortality and the severity of cervical spinal cord injury. According to Higashi T. et al., there was no discernible relationship between the degree of neurological impairment and mortality risk (20). On the other hand, according to Shao et al., patients with cervical SCI who have upper-level cervical spinal cord injury are at risk for early mortality (19). In agreement with the later study, the results of this study state that the level of neurological impairment was a significant risk factor for mortality. The discrepancy with the result of the earlier study might be due to the difference in the study population, sample size, methodology, the age of the participant, and other factors.

According to research by Ali R.et al., neurological deficits, comorbidities, and the presence of other injuries were significant predictive factors for complications and death in patients with cervical spine injuries (P < 0.05). The most common concomitant injuries were long bone fractures (11.6%), rib fractures (11.1%), and head injuries (7.6%) (35). In line with this study, 9.4% of patients in our study had extremities injuries, which were linked to death (P value < 0.05).

Katherine E. et al., categorized patients into two groups based on the length of their inpatient hospital stay: extended (E-LOS; >75^th^ percentile) and normal (N-LOS; <75^th^ percentile) (36). Additionally, Kazuyoshi K. et al., classified patients as having a normal length of stay (LOS) (<75^th^ percentile LOS) or a prolonged LOS (>75^th^ percentile LOS) in their multicenter study (37). Longer hospital stays (LOSs) have been linked to a number of unfavorable outcomes. In this study, 45 patients (30.2%) stayed longer than the 75^th^ percentile, which is 15 days. It has been demonstrated that longer LOS increases healthcare expenses and resource use. Advanced patient age, emergency admission, ASIA grade, operational management, and the occurrence of one or more related injuries were factors that were substantially linked to extended length of stay (LOS) (14). Consistent with the aforementioned research, a longer hospital stay was substantially linked to both neurosurgical intervention and the existence of a concomitant chest injury.

### Limitations of the study

All the limitations associated with a retrospective study also apply to this one, including the requirement to account for confounding variables. Only in-hospital mortality was evaluated in this study; long-term mortality was not looked at. Factors such as the patient’s previous comorbidity, length of mechanical ventilation in the intensive care unit, and requirement for a tracheostomy, of which have an impact on mortality in patients with cervical spine injuries. These are not taken into account in this analysis. Additionally, the research excludes institutional, social, and personal elements that may have an impact on the length of hospital stay.

Ultimately, because the study was only carried out at one location with a small sample size, it’s possible that the results cannot be applied to other contexts.

## Conclusion

This study showed the common cause (mechanism) was RTA and C7 was the common injury level. C3 injury level, ASIA A neurologic deficit, and having associated injury were associated with mortality. Undergoing neurosurgical intervention and the presence of associated injury were associated with prolonged length of hospital stay (PLOS). In the presence of the above limitation, the aforementioned findings should facilitate the development of preventative policies and their implementation by the relevant stakeholders in the health and transportation sectors.

## Data Availability

We all are ready to submit any additional files up on request

## Acknowledgment

We would like to thank St. Paul’s Hospital Millennium Medical College and the Department of Emergency and Critical Care Medicine for the opportunity to conduct this research and financial support.

